# Safety and immunogenicity of a quadrivalent meningococcal polysaccharide vaccine (MPV ACYW135) compared with a quadrivalent meningococcal conjugate vaccine (Menactra^®^): a study protocol of a randomized non-inferiority trial in Mali

**DOI:** 10.1101/2025.11.28.25341186

**Authors:** Simonetta Viviani, Ruoyu Hu, Samba O Sow, Milagritos D Tapia, Fadima Cheick Haidara, Fatoumata Diallo, Youssouf Traore, Awa Traoré, Mamoudou Kodio, Lei Shi

## Abstract

A phase IV clinical trial is designed to compare the safety and immunogenicity of a quadrivalent meningococcal polysaccharide vaccine (Walvax MPV ACYW135) compared with a quadrivalent meningococcal conjugate vaccine (Sanofi Pasteur Menactra^®^) in children aged 2-10 years in Mali, a country within the meningitis belt, to demonstrate the benefit that might be brought by vaccinations with meningococcal polysaccharide vaccines. Eligible subjects will be randomized at a 1:1 ratio to receive either Walvax MPV ACYW135 or Sanofi Pasteur Menactra^®^. All subjects will be followed up for solicited/unsolicited adverse events and any serious adverse events. Immunogenicity will be determined with the level (titer) of functional antibodies by serum bactericidal assay using baby rabbit complement (rSBA) with non-inferiority demonstrated.

**Clinical Trial Registration:** NCT04450498

## Introduction

Meningococcal diseases are a series of diseases caused by the infection with *Neisseria meningitidis* (*N. meningitidis*), a fastidious, encapsulated, aerobic gram-negative diplococcus. Meningococcal diseases pose significant morbidity and mortality in children and young adults worldwide through epidemic or sporadic meningitis and/or septicemia. According to the World Health Organization (WHO), explosive epidemics with incidence rates of up to 1000 cases per 100,000 inhabitants have been reported, particularly in sub-Saharan Africa^1^, where the so called “meningitis belt” is located, with the highest burden of meningococcal diseases and at high risk of recurrent epidemics of meningococcal meningitis.

At least 12 serogroups based on unique capsular polysaccharides of *N. meningitidis*, with serogroups A, B, C, W, X, and Y responsible for the majority of meningococcal infections^2^. Effective vaccines are available against 5 of 6 disease-causing serogroups. WHO recommends that countries of the African meningitis belt to conduct preventive campaigns with appropriate meningococcal vaccines in specified populations, as well as to introduce meningococcal vaccines into the routine immunization programme within 1-5 years^1^. In countries where the disease occurs less frequently (< 2 cases per 100,000 population/year), meningococcal vaccination is also recommended for defined risk groups.

Despite successful antimicrobial treatments for the past 70–80 years, there is now a high prevalence of *N. meningitidis* strains with resistance to most commonly used antimicrobials^3^. The development and introduction of effective vaccines against different meningococcal serogroups poses great impact on the global epidemiology of meningococcal diseases. The majority of observed changes in the epidemiological patterns of invasive meningococcal diseases (IMDs) have been reported to occur following the implementation of vaccines directed against *N. meningitidis* serogroups C and A. Importantly, these vaccines have been primarily employed in Western countries and sub-Saharan Africa. The introduction of novel vaccines against additional serogroups is expected to exert a further impact on reducing the disease burden. Furthermore, at least two subcapsular protein-based serogroup B meningococcal vaccines, whose application is predicted to increase, are likely to confer cross-protective immunity against other meningococcal strains^4^.

Menactra^®^ (Meningococcal [Groups A, C, Y and W-135] Polysaccharide Diphtheria Toxoid Conjugate Vaccine, Sanofi Pasteur Inc., Swiftwater, PA, USA) was approved by the Food and Drug Administration in 2005 and became available for public use. Menactra^®^ was the first meningococcal conjugate vaccine and received FDA approval on the basis that the vaccine was not inferior in safety or immunogenicity when compared with Menomune^®^, the only FDA approved meningococcal vaccine available at the time. Menactra^®^ has been widely used since its licensure with solid evidence of its immunogenicity, safety and effectiveness among individuals aged 9 months through 55 years.

A Group ACYW135 Meningococcal Polysaccharide Vaccine (MPV ACYW135) developed by Yuxi Walvax Biotechnology Co., Ltd. (Walvax, Yuxi, Yunnan, China) has been widely used in China since 2012. This article discusses a study protocol designed to compare the safety and immunogenicity of Walvax MPV ACYW135 as compared to Sanofi Pasteur Menactra^®^ in children aged 2-10 years in Mali, a country within the meningitis belt, to demonstrate the benefit that might be brought by vaccinations with meningococcal polysaccharide vaccines.

## Methods

### Overall Study Design

The study is designed as a Phase IV, randomized, observer-blind, controlled study to be conducted at the Centre pour le Développement des Vaccins du Mali (CVD-Mali), in Bamako, Mali, among children of 2 to 10 years. Eligible subjects will be randomized at a 1:1 ratio to receive a single intramuscular injection of Walvax MPV ACYW135 or Sanofi Pasteur Menactra^®^ with 130 subjects planned to be included in each arm (overall sample size of 260). All subjects will be followed up for local and systemic solicited AEs for 6 days after the day of vaccination, unsolicited AEs for 30 days, and SAEs for 180 (±15) days, or approximately 6 months after vaccination. All subjects will be evaluated for immunogenicity at baseline and 1 month post vaccination. Please see Error! Reference source not found. for an overview of the overall study design.

**Figure 1.**
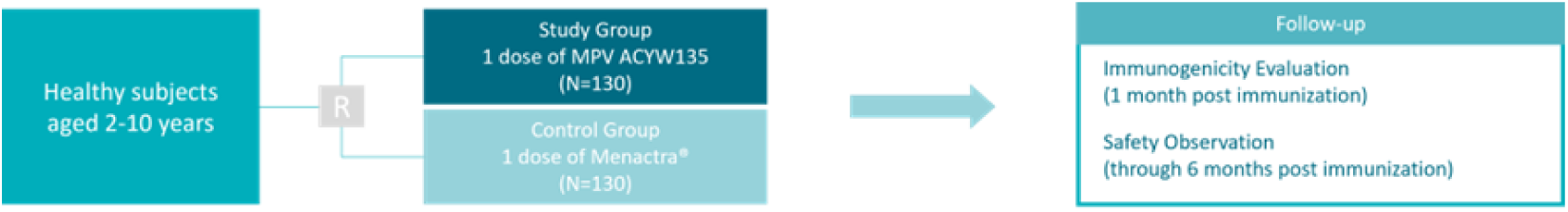
Diagram of Study Procedures

### Study Objectives and Outcomes

#### Primary Objective

To demonstrate non-inferior immunogenicity of one dose of Walvax MPV ACYW135 vaccine as compared to one dose of Sanofi Pasteur Menactra^®^ in subjects aged 2-10 years (both included).

#### Secondary Objective

To assess the safety of one dose of Walvax MPV ACYW135 as compared to one dose of Sanofi Pasteur Menactra^®^.

### Primary Immunogenicity Endpoint (Non-inferiority)

#### Antibody response at Day 30 post vaccination

- Percentage of subjects with rSBA (Serum Bactericidal Activity using baby rabbit complement) titer ≥1:128 to groups A, C, Y, and W135 meningococcal capsular polysaccharide in both vaccine groups 30 days after immunization.

### Secondary Immunogenicity Endpoints

#### Immunogenicity endpoints: at Day 30 post vaccination

- rSBA Geometric mean antibody titers to groups A, C, Y, and W135 meningococcal capsular polysaccharide in both vaccine groups.
- Percentage of subjects with rSBA titer ≥1:8 to groups A, C, Y, and W135 meningococcal capsular polysaccharide in both vaccine groups 30 days after immunization.
- Seroconversion rates as defined by proportion of subjects with ≥ 4-fold increase 30 days after immunization with respect to baseline of rSBA antibodies to groups A, C, Y, and W135 meningococcal capsular polysaccharide in both vaccine groups 30 days after immunization.
- Seroconversion rates as defined by proportion of subjects with ≥ 2-fold increase 30 days after immunization with respect to baseline of rSBA antibodies to groups A, C, Y, and W135 meningococcal capsular polysaccharide in both vaccine groups.

### Secondary Safety Endpoints

#### Safety endpoints: at Day 30 post vaccination and until end of study (Day 180 ± 15 days approximately six months)

- Percentages of subjects with post-immunization local and systemic solicited AEs within 7 days following vaccination in both vaccine groups
- Percentages of subjects with reported AEs within 30 days following vaccination in both vaccine groups
- Percentages of subjects with reported SAEs within 6 months following vaccination in both vaccine groups

### Eligibility criteria

#### Inclusion Criteria

A subject was eligible for inclusion if ALL of the following applied at the time of enrolment:

1. Age 2 to 10 years of age (both included).
2. Written informed consent obtained from the mother, father, or guardian of the child.
3. Free of obvious health problems as established by medical history including physical examination and clinical judgment of the investigator.
4. Mother, father, or guardian capable and willing to bring their child or to receive home visits for their child for all follow-up visits.
5. Residence in the study area during the study period.

#### Exclusion Criteria

Subjects with any of the following criteria at the time of study entry were not eligible for participation:

1. Vaccination against groups A, C, Y, W Neisseria meningitidis in the previous 3 months
2. History of allergic disease or known hypersensitivity to any component of the two study vaccines.
3. History of serious adverse reactions following administration of vaccines included in the local program of immunization.
4. Administration of any other vaccine within 30 days prior to administration of study vaccines or planned vaccination during the first four weeks after the study vaccination.
5. Use of any investigational or nonregistered product within 60 days prior to the administration of study vaccines.
6. Administration of immunoglobulins and/or any blood products or planned administration during the study participation period.
7. Chronic administration (defined as more than 14 days) of immunosuppressants or other immune-modifying agents since birth (including systemic or inhaled corticosteroids, this means prednisone, or equivalent, ≥0.5 mg/kg/day; topical steroids are allowed).
8. A family history of congenital or hereditary immunodeficiency.
9. History of meningitis or seizures, or any neurological disorder, convulsions, or active tuberculosis.
10. Major congenital defects or serious chronic illness, including malnutrition (i.e., weight less than or equal to 3 standard deviations below the mean for 2-5 years old) and immunodeficiency disorder (as per investigator’s judgment)
11. Acute disease at the time of enrolment (acute disease being defined as the presence of a moderate or severe illness with or without fever) resulting in a temporary exclusion.
12. Acute or chronic, clinically significant pulmonary, cardiovascular, hepatic, or renal functional abnormality, as determined by medical history, physical examination, or laboratory tests, which in the opinion of the investigator might interfere with the well-being of the subject study objectives.
13. Any condition or criterion that in the opinion of the investigator might compromise the well-being of the subject or the compliance with study procedures or interfere with the outcome of the study

### Ethics

Regulatory notification for this study will be sent to responsible authorities according to the applicable country-specific laws and regulations. Approval will be obtained from the appropriate regulatory authority prior to initiation of the study. The study will be conducted according to *Good Clinical Practice* (ICH guidelines)^5^, the Declaration of Helsinki, and applicable laws and regulations of the country where the study takes place. The protocol, along with other related documents including the Informed Consent Form and Diary Card used for the study, will be reviewed and approved by the Independent Ethics Committee (IEC) responsible for the corresponding study site.

The investigator or his/her designee will inform the subject’s parents/legal guardian of all aspects pertaining to the subject’s participation in the study. The process for obtaining subject informed consent will be in accordance with all applicable regulatory requirements. Prior to including any subject in the clinical study, subject’s parents/ guardian freely expressed informed consent must be obtained in writing. The written informed consent must be signed and dated by investigator or his/her designee, the subject’s parents/ guardians prior to any study related procedure.

### Intervention and Group Assignment

As part of the clinical development, a non-inferiority clinical trial to demonstrate non-inferior immunogenicity as compared to a marketed similar vaccine was conducted in China during 2007-2008. At that time, the only available similar licensed vaccine in China was the meningococcal groups A and C polysaccharide vaccine manufactured by Lanzhou Institute of Biological Product Co., Ltd. However, a formal non-inferior immunogenicity study of Walvax MPV ACYW135 as compared to a globally licensed similar vaccine had not been conducted. A post-marketing study to demonstrate non-inferior immunogenicity as compared to a globally licensed meningococcal polysaccharide ACYW vaccine is part of the commitment to comply with the requirements of WHO pre-qualification team. While planning the study it became evident that production of the only two globally licensed and prequalified polysaccharide vaccines Menomune^®^ (manufactured by Sanofi Pasteur) and Mencevax^®^ (manufactured by GSK, acquired by Pfizer^6^) were discontinued^7,8^ by the respective company, and no other ACYW polysaccharide vaccine is available in the market to be purchased and used as the comparator for this study. Therefore, the WHO-prequalified conjugate meningococcal ACYW vaccine Sanofi Pasteur Menactra^®^ was chosen as the comparator vaccine in this non-inferiority study.

### Walvax MPV ACYW135^9^

MPV ACYW135 is a lyophilized vaccine of purified meningococcal capsular polysaccharides of group A, C, Y and W135 *Neisseria meningitides* (*N. meningitides*).

MPV ACYW135 is provided as a single human dose with a vial of white loose sterile powder and another vial of diluent. The lyophilized polysaccharide vaccine should be reconstituted immediately before use with the accompanying diluent (sterile water for injection). After reconstitution with diluent, the vaccine shall immediately turn into a clear, colorless solution. MPV ACYW135 shall be injected subcutaneously after reconstitution.

MPV ACYW135 is supplied as a single dose vial of lyophilized powder, with corresponding single dose of diluent. Each dose of 0.5 mL contains:

|  |  |  |
| --- | --- | --- |
| **Active ingredients:** |  |  |
| Polysaccharide for serogroup A, C, Y and W135 |  | 50 µg each |
| **Excipients:** | Lactose | 4 mg |
|  | Sodium chloride | 4.25 mg |
| **Diluent:** | Sterile water for injection | 0.5 mL |

### Sanofi Pasteur Menactra^®10^

Menactra^®^ is a sterile, clear to slightly turbid liquid of *Neisseria meningitidis* serogroups A, C, Y and W-135 capsular polysaccharide antigens individually conjugated to diphtheria toxoid protein. The polysaccharides are covalently linked to diphtheria toxoid and purified by serial diafiltration. The four meningococcal components, present as individual serogroup-specific glycoconjugates, compose the final formulated vaccine. Menactra^®^ should be injected by the intramuscular route.

Menactra^®^ is a clear to slightly turbid liquid. Each single dose (0.5 mL) is formulated to contain:

#### Active Ingredients

4 μg each of meningococcal A, C, Y and W-135 polysaccharides conjugated to a total of approximately 48 μg of a diphtheria toxoid protein carrier

|  |  |
| --- | --- |
| Sodium Chloride | 4.25 mg |
| Sodium Phosphate, Dibasic, Anhydrous | QS phosphate 10 mM |
| Sodium Phosphate, Monobasic | QS phosphate 10 mM |

#### Other Ingredients

Each participant will receive a single intramuscular injection of one of the two vaccines either Walvax MPV ACYW135 or Sanofi Pasteur Menactra^®^ according to the vaccine group assignment and will be followed up for one month for immunogenicity evaluation and for 6 months for safety evaluation. Please see **Table 1** for the assignment of subjects in each group.

*Other Ingredients*
|  |  |
| --- | --- |
| Water for Injection | QS 0.5 mL |

**Table 1.** Assignment of Subjects.

| Group | Intervention | Dose Regimen | Sample Size |
| --- | --- | --- | --- |
| Study Group | Walvax MPV<br>ACYW135 | Single dose | 130 |
| Control Group | Sanofi Pasteur<br>Menactra® | Single dose | 130 |

#### Schedule of Visits

All subject’s parents/guardians will be asked to attend the study site for a total of 4 visits (Visit 1 to Visit 4). At Visit 1 (Day 0), potential participants whose parents/legal guardians have the informed consent signed will be screened for general health status and those who fulfil all inclusion without any exclusion criteria met will be enrolled into the study. Eligible subjects will be given with 1 dose of either Walvax MPV ACYW135 or Sanofi Pasteur Menactra^®^ according to the vaccine group assignment after randomization. Subjects will be observed for 30 minutes at study site for any immediate post-immunization reactions after vaccination. Within 6 consecutive days post vaccination, reactogenicity and safety will be assessed daily through home visits performed by local site field staffs, during which solicited local and systemic solicited AEs or any other AEs will be collected. At Visit 2 (Day 7 +2 days), a medical evaluation will be carried out for the subject either at home or at the study site. With the period between Visit 2 (Day 7 +2 days) and Visit 3 (Day 30 +7 days), parents/legally acceptable guardians will be instructed to report to study site any health problems that their children encountered. All AEs will be collected and monitored for 30 days after vaccination (until Visit 3); thereafter, only SAEs will be monitored reported throughout the entire study duration for approximately 6 months. The end-of-study visit, Visit 4 (Day 180 ± 15 days), will be performed by medically qualified staffs either at home or at the study site. Information collected within 6 days after immunization and until Day 30 will be entered into the electronic Case Report Form (eCRF).

Blood samples will be collected from all subjects at Visit 1 (Day 0) for baseline values and at Visit 3 (Day 30 + 7 days) after vaccination to evaluate the immune response to study vaccines.

Please see **Table 2** for detailed study procedures performed.

**Table 2.**
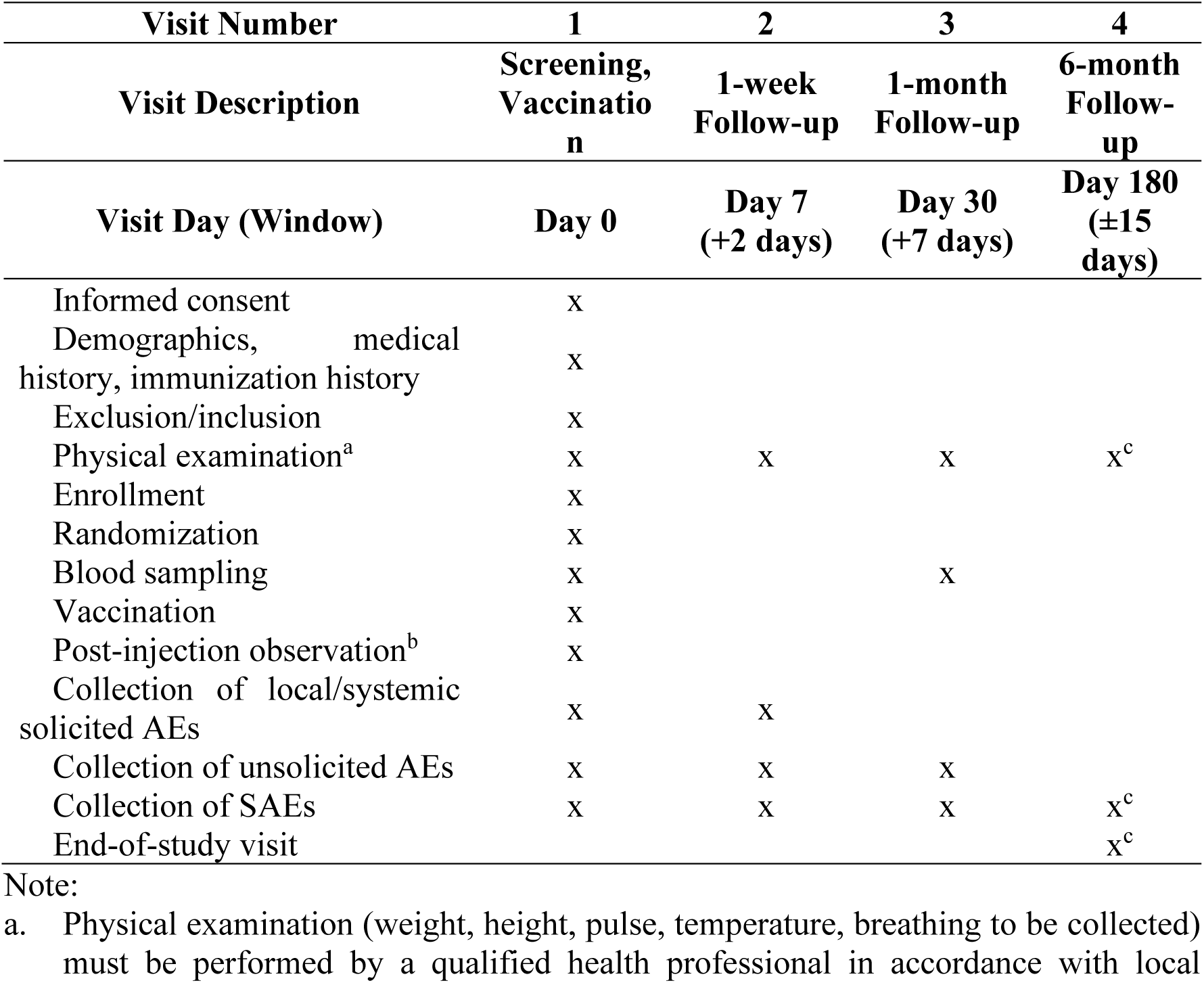
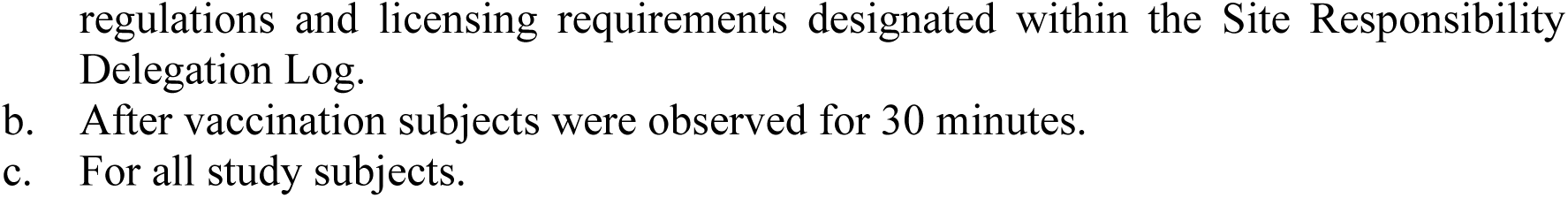
Schedule of Visits.

| **Visit Number** | **1** | **2** | **3** | **4** |
| --- | --- | --- | --- | --- |
| **Visit Description** | **Screening, Vaccination** | **1-week Follow-up** | **1-month Follow-up** | **6-month Follow-up** |
| **Visit Day (Window)** | **Day 0** | **Day 7 (+2 days)** | **Day 30 (+7 days)** | **Day 180 (±15 days)** |
| Informed consent | x |  |  |  |
| Demographics, medical history, immunization history | x |  |  |  |
| Exclusion/inclusion | x |  |  |  |
| Physical examination<sup>a</sup> | x | x | x | x<sup>c</sup> |
| Enrollment | x |  |  |  |
| Randomization | x |  |  |  |
| Blood sampling | x |  | x |  |
| Vaccination | x |  |  |  |
| Post-injection observation<sup>b</sup> | x |  |  |  |
| Collection of local/systemic solicited AEs | x | x |  |  |
| Collection of unsolicited AEs | x | x | x |  |
| Collection of SAEs | x | x | x | x<sup>c</sup> |
| End-of-study visit |  |  |  | x<sup>c</sup> |

Note:
- a. Physical examination (weight, height, pulse, temperature, breathing to be collected) must be performed by a qualified health professional in accordance with local
regulations and licensing requirements designated within the Site Responsibility Delegation Log.
- b. After vaccination subjects were observed for 30 minutes. - c. For all study subjects.

### Endpoint Assessment

#### Safety Assessment

After administration of the allocated vaccine, the subject will be monitored for 30 minutes, with appropriate medical treatment readily available in case of any anaphylactic reaction. Blood pressure and pulse rate will be measured according to investigator’s medical judgment. At Day 0, after vaccination, subject’s parents/ guardians will be informed that on the subsequent 6 days site field staff will visit home on a daily basis for collection of local and systemic solicited AEs and any other AEs. In addition, the subject’s parents/ guardians will be asked to report any AEs whether or not he/she believes it is related to the study vaccination. During Visit 2 at Day 7 (+2) medically qualified staff will perform a medical examination of study subjects either at home or at the study clinic. Investigators will reconcile and transcribe all information on post-immunization local and systemic reactions, all other adverse events and medical examination findings into the appropriate eCRF pages, using medical language. Local and systemic solicited AEs to be collected and reported by site field staff consist of: injection site induration and injection site pain, irritability, loss of appetite, drowsiness, vomiting and fever. The severity and causal relationship to investigational vaccines will be determined by the investigators in accordance with study requirements.

After Visit 2 (Day 7+2) subject’s parents or guardians will be instructed to report to investigator any adverse event that may occur whether or not perceived as related to study vaccine. At Visit 3 (Day 30 +15) investigator will reconcile, transcribe any reported adverse event, concomitant medication and medical examination findings in the appropriate eCRF. All SAEs will be collected and followed up for six months (180 days +15 after vaccination.

#### Immunogenicity Assessment

Approximately 5 mL of whole blood will be taken from all subjects at Day 0 (Visit 1) just before vaccination (baseline) and at Day 30 (+7) (Visit 3) after vaccination. Serum samples will be stored frozen at −20°C at study site and shipped to Sponsor designated laboratories to perform immunogenicity analyses. A validated serum bactericidal assay using baby rabbit complement (rSBA) will be used to measure the level (titer) of functional antibody in human sera to *Neisseria meningitidis* group A, C, Y, W135 capsular polysaccharide. The measurements will be performed at the Vaccine Evaluation Department, Health Protection Agency (HPA), Manchester, U.K.

#### Randomization and Blinding

For vaccine allocation, a randomization blocking scheme (1:1) will be used so as to ensure that balance between vaccines will be maintained. A randomization number will identify the vaccine to be administered to each subject. Subjects will be randomized sequentially in the order they are enrolled.

This trial was carried out in an observer-blind fashion and blinding will be maintained until database was locked with data collected until Day 30 (Visit 3) after vaccination. At study site, the principal investigator will assign unblinded designated person(s) responsible for vaccine handling, administration (“Vaccinator”) and accountability to ensure that blinding will be protected throughout the study procedures and that no other site personnel involved in the conduct of the study had access to information regarding blinding. The randomization code is only to be broken in case of a medical event, which, in the opinion of the investigator, couldn’t be treated without knowing the nature of the vaccine. Any emergency unblinding is required to be documented.

### Statistical Analyses

#### Estimation of sample size

The sample size was calculated based on non-inferiority test with alpha level of 0.025 (one-sided) and 80% power, assuming seroconversion rate in control group was 95% with non-inferiority margin at 10%. The minimal sample size required for the study is 124 per arm. After adjusting for 5% drop-out, the final sample size required is 130 per arm.

#### Statistical Hypothesis

*H0*: Seroconversion rate of study group is inferior to that of control group;

*HA*: Seroconversion rate of study group is non-inferior to that of control group.

## Overall Statistical Methods

Baseline characteristics of subjects in the two vaccines groups will be compared, using *chi-square* test or Fisher’s exact test for categorical variables and Student’s *t* test for continuous variables respectively. Within each vaccine group, for each serogroup, the seroconversion rate together with the two-sided 95% CI will be calculated; for each antibody titer, the geometric mean antibody titer and the two-sided 95% Cl at baseline and at 30-day after vaccination will be calculated. Difference of seroconversion rates between groups with two-sided 95% CI will be determined. Difference of geometric mean antibody titers between groups will be compared using *t* test. AEs will be described and compared between two groups using *chi-square* test or Fisher’s exact test. Acceptance difference of seroconversion rate at ≤ 10% for the upper bound of the 95% CI will be considered for non-inferiority.

No missing data imputation techniques will be planned for the primary analysis of this study. No Independent Data Monitoring Committee (IDMC) was convened in this study. No interim analysis will be performed in this study.

## Data Availability

All data produced in the present study are available upon reasonable request to the authors

## Acknowledgements

The authors thank the children and their families for participating in this study with prior consent. The authors gratefully acknowledge investigators at the Centre pour le Développement des Vaccins du Mali (CVD-Mali) for their contribution to performing this study.

## Ethical Approval Statement

Regulatory notification for this study will be sent to responsible authorities according to the applicable country-specific laws and regulations. Approval will be obtained from the appropriate regulatory authority prior to initiation of the study. The study will be conducted according to Good Clinical Practice (ICH guidelines), the Declaration of Helsinki, and applicable laws and regulations of the country where the study takes place. The protocol, along with other related documents including the Informed Consent Form and Diary Card used for the study, will be reviewed and approved by the Independent Ethics Committee (IEC) responsible for the corresponding study site.

The investigator or his/her designee will inform the subject’s parents/legal guardian of all aspects pertaining to the subject’s participation in the study. The process for obtaining subject informed consent will be in accordance with all applicable regulatory requirements. Prior to including any subject in the clinical study, subject’s parents/ guardian freely expressed informed consent must be obtained in writing. The written informed consent must be signed and dated by investigator or his/her designee, the subject’s parents/ guardians prior to any study related procedure. The study was registered on *<u>clinicaltrials.gov</u>* as NCT04450498.

## Author Contribution

SV designed the study. SS, MDT, FCH, FD, YT, AT and MK coordinated and implemented the study. RH and LS wrote the first draft manuscript. All the other authors reviewed and contributed to the final version.

